# Drug repositioning for immunotherapy in breast cancer using single-cell and spatial transcriptomics analysis

**DOI:** 10.1101/2022.11.18.22282496

**Authors:** Elyas Mohammadi, Han Jin, Cheng Zhang, Neda Shafizade, Samira Dashty, Simon Lam, Mojtaba Tahmoorespur, Adil Mardinoglu, Mohammad Hadi Sekhavati

## Abstract

**Background:** Immunomodulatory peptides are capable of stimulating or suppressing the immune system. Hence, deregulation of them can be considered as an immunotherapy approach. These peptides may have dual behavior in response to different infections. For instance, an antimicrobial peptide may act as an anticancer, tumor marker or even cause cancer progression.

**Methods:** In this study, we used single-cell RNA sequencing and spatial transcriptomiocs analysis to investigate the deregulation of immunomodulatory peptides in malignant versus normal human breast epithelial cells. We validated the obtained results in chromatin accessibility level. Furthermore, we used a drug repositioning approach to change the expression of these peptides based on their role in cancer biology.

**Results:** Ten immunomodulatory peptides were upregulated in breast cancer versus normal. Chromatin was more accessible for these peptides in cancer cell lines versus normal. Among these ten peptides, five of them were tumor drivers (i.e., BST2, GAPDH, S100A8, S100A9 and HMGB1), three of them were anticancer (i.e., H2AFJ, SCGB2A1 and HMGN2), S100A7 had dual behavior in different cancers and ZG16B was a tumor marker. Using the LINCS L1000 database, we proposed a list of drugs that can deregulate the candidate peptides according to their role in the progression of malignancy.

**Conclusions:** Immunomodulatory peptides can be considered as drug targets based on their role in cancer biology.

## Introduction

Breast cancer (BC), in general, has low immunogenicity [1]; however, immune activation can be pivotal in BC prognosis and treatment [2]. Cancer immunotherapy, also known as immuno-oncology, is a form of cancer treatment that educates, boosts or inhibits the immune system components to prevent, control, or eliminate cancer [3]. Antimicrobial and immunomodulatory peptides (AMPs) are a part of the innate immune system [4]. AMPs may directly interact with pathogens and/or malignant cells or indirectly modulate the immune system leading to the elimination of target cells [4, 5]. On the other hand, AMPs that are capable of eliminating pathogens may promote cancer progression [6]. Hence, the knowledge of the cancer-specific response of AMPs and finding ways to improve or suppress it can pave the way for immunotherapy in precision medicine.

To better understand the immunotherapy targets, various techniques are deployed to assess gene expression [7] including conventional bulk transcriptomics and single-cell transcriptomics (scRNA-seq) [8]. Bulk transcriptomics measures the average of gene expression across all cells. scRNA-seq deals with gene counts in each cell; hence, it provides the opportunity to categorize and annotate the cell types based on their expression profiles. Breast cancer is heterogeneous and also comprises—beyond malignant cells— stromal and immune tumor-associated cells. Although bulk transcriptomics still has its own advantages in cancer immunotherapy [8], for precision onco-immunotherapy scRNA-seq provides higher resolution for both malignant and immune cell profiling [8, 9].

Single-cell studies have improved drug target and biomarker discovery in immunotherapy [10-12], while finding the potential drugs for these targets is still challenging [13]. Drug repositioning against immunotherapy targets explores the therapeutic use of existing clinically approved, off-patent drugs. These chemicals have known modes of action and targets for another indication. Thus, exploring them minimizes the cost of therapy, time and risk [14-17].

Deregulation of immunomodulatory peptides is considered as an independent immunotherapy treatment or auxiliary to obtain better results through combination therapy [18, 19]. In this study, we aimed to investigate the endogenous expression of, to the best of our knowledge, all so far known human antimicrobial and immunomodulatory peptides in BC [6]. To do so, we utilized BC and normal breast scRNA-seq data in addition to Assay for Transposase-Accessible Chromatin using sequencing (ATAC-seq) data from BC cell lines. This in turn improved the resolution of our analysis as we could extract and compare malignant versus nonmalignant epithelial cells at different omics levels (i.e., chromatin and gene levels). Next, according to the role of upregulated immunomodulatory peptides in cancer biology, we proposed drugs that can deregulate them accordingly.

## Methods

### Data collection and filtering

The processed data for 26 BC patients was obtained from the study conducted by wu et al. 2021 [1]. Cells were previously filtered based on percentage of mitochondrial genes and high and low number of features [1]. Out of the 26 patients, 20 of them whose epithelial cells could be distinguished as normal or malignant were selected for downstream analysis. The classification of epithelial cells in these patients was previously performed using the InferCNV R package [20]. InferCNV can identify evidence for large-scale chromosomal copy number variations by exploring expression intensity of genes across positions of the genome in comparison to the average or a set of reference normal cells [20]. The processed data for 13 normal breast samples was retrieved from the study conducted by Pal et al 2021 [21]. The percentage of mitochondrial genes in each cell was calculated using PercentageFeatureSet from Seurat package (Version 4.1.0) [22]. According to Pal et al. 2021, no more than 20% mitochondrial reads were generally allowed per cell, although the upper limit was increased as high as 40% for a small number of libraries. Cells with exceptionally high numbers of reads or genes detected were also filtered to minimize the occurrence of doublets. An average of 5000 cells per sample remained after this quality filtering.

Normalized chromatin accessibility data for four BC cell lines were retrieved. These cells included two TNBC cell lines, i.e MDA_MB_231 and MDA_MB_436 with one repetition [23], MCF7 as an ER BC cell line with a repetition [24], and MCF10A as a normal breast epithelial cell.

Transcriptomics data was processed in the Seurat R package (Version 4.1.0). The top 6000 features by variance across the datasets were selected using variance-stabilizing transform (VST) method and scaled using the default parameters of the ScaleData function of Seurat. Finally, dimensionality reduction was performed based on these scaled features and top 15 dimensions using default parameters of RunPCA and RunUMAP functions of the Seurat [22]. We used a Shared Nearest-neighbor (SNN) graph construction method using FindNeighbors function in addition to a modularity optimization of SNN results using FindClusters function of the Seurat R package with default parameters to do an unsupervised clustering and categorize the normal breast single cell dataset.

### Cell Annotation

The BC dataset was previously annotated. To annotate the single cells of normal breast samples as query set we used SingleR package (Version 1.8.1) and Human Primary Cell Atlas (HPCA) [25] and Blueprint Epigenomics (BP) [26] as two previously annotated references and processed them in Seurat. HPCA and BP references were merged and their labels were transferred to the query dataset. To illustrate the annotation results, PlotScoreHeatmap of the SingleR R package shows the assigned label scores to each cell. The package pheatmap (version 1.0.12) was used to depict the consistency between unsupervised clustering and supervised cell annotation. PlotDeltaDistribution plots the distribution of delta values for each cell, i.e., the difference between the score for the assigned label and the median across all labels for each cell. Finally, the results from all these steps were transferred to the defined clusters of normal breast samples.

### Data integration

Normal and malignant epithelial cells were extracted from normal and BC datasets and then merged, normalized using the LogNormalization method. Next, top 10000 variable features between two datasets were defined by the FindIntegrationFeatures function for data integration. Based on these variable genes we used the FindIntegrationAnchors function to determine the cell pairwise correspondences across single cells datasets [27]. Finally, the IntegrateData was used to perform data integration by pre-computed anchor set. For the validation purposes, the same pipeline was applied to the SCTransform normalization method [28] as well and the results after differential gene expression were compared between two normalization methods.

### Differential gene expression

Differential gene expression analysis between malignant and normal epithelial cells was done on both integrated objects which were normalized through LogNormaization and SCTransform methods. DEGs were defined using FindMarkers function by pairwise comparison of BC subtypes and normal epithelial cells, separately. Next, all so far known immunomodulatory peptides were extracted from University of Debrecen Antimicrobial and Immunomodulatory Peptide (UDAMP) database [6]. Deregulation of these genes were investigated among DEGs and illustrated by EnhancedVolcano plot (Version 1.12.0). Afterwards, deregulated AMPs were classified based on their role in BC biology i.e the genes that are in the favor of cancer progression and should be downregulated (Down), the genes associated with positive outcome and cancer elimination (Up) and the genes that are BC markers or have unclear behavior (No change). Next, the average of gene expression for these three categories were computed using the AverageExpression function.

### Gene-Gene correlation and functional analysis

We performed Pearson gene-gene correlation for the candidate AMPs among top 10000 variable genes of integrated object. The p-values for the correlations were adjusted using the False Discovery Rate (FDR) method. Only positive correlations with adjusted p-value lower than 0.05 were extracted. Next, top 20 gene correlations for each AMP were applied to Gene Ontology (GO) and KEGG pathway analysis using ClusterProfiler (Version 4.2.2) and Benjamini Hochberg (BH) p-value adjusting method. The R package org.Hs.eg.db (Version 3.14.0) representing the human genome was utilized for GO analysis and the ensemble based annotation package EnsDb.Hsapiens.v86 (Version 2.99.0) built based on GRCh38 was used for KEGG analysis.

### Spatial transcriptomics data analysis

ST profiles of four different tissue sections were extracted from the study conducted by Wu et al. (2021). All the gene count matrixes and images were imported seperately using Read10X and Read10X_Image Seurat functions, respectively. Images and gene counts were later merged using CreateSeuratObject function of Seurat R package. In order to deconvolute the spatial voxels, we performed the ST and scRNA-seq data integration using the profiles from the same patients. To do so, after data normalization using SCTransfrom function, we determined the anchor set between reference (i.e, scRNA-seq datasets) and query (i.e, ST profiles) using FindTransferAnchors function of Seurat R package. Using anchors, one can transfer information between two sets. Afterwards, we transferred cell type informations from scRNA-seq to ST datasets using TransferData function of Seurat R package.

### Drug repositioning

The metadata for all BC cell lines in LINCS L1000 database were extracted and categorized based on BC subtypes i.e, TNBC, HER2 and ER. Number of available perturbagens with 10 μm dose and 24 hours treatment duration for each BC cell line was investigated by the Slinky R package (Version 1.12.0). The BC cell line with the most number of perturbagens based on aforementioned criteria was selected for downstream analysis. The control expression profiles were considered as treated with DMSO with the same dose and duration criteria. Next, the LFCs for the effect of each drug on the expression profile of the selected BC cell line was calculated. To do so, first the mean of gene expression for the effect of each drug across the repetitions was calculated for both control and treatment. Next, we computed the log2 of the mean of gene expressions and finally, the log transformed results for control and treatments were subtracted.

The LFCs of the candidate AMPs across all drugs were extracted from the log fold change matrix. The profiles were categorized to the drug repositioning purposes based on the role of these genes in cancer biology i.e, Down (downregulation drug repositioning strategy) and Up (upregulation drug repositioning strategy). For the genes that should be up or downregulated through drug repositioning, the profiles were sorted descending and ascending, respectively. Hence, top 20 drugs that upregulate or downregulate the genes were chosen based on the purpose of drug repositioning. Finally, the effect of the proposed drugs on deregulation of genes were plotted.

## Results

### Extracting the transcriptomics profiles of epithelial cells

In this study, we aimed to investigate the deregulation of antimicrobial and immunomodulatory peptides in malignant versus nonmalignant human breast epithelial cells at different omics levels. Hence, we would be able to predict the potential candidate drugs that can change the expression of these peptides in favor of patients (Figure 1). We retrieved two publicly available scRNA-seq datasets (details in Data Availability) from 13 normal [21] and 26 BC patients [29] (5 HER2, 11 ER and 10 TNBC subtypes). The cells in the BC dataset were previously annotated for cell types. For 20 patients (3 HER2, 9 ER and 8 TNBC subtypes) in the BC dataset, the epithelial cells, as the most common site for the development of BC [30], were categorized into malignant and nonmalignant cells. The classification of epithelial cells was done by identifying evidence for large-scale chromosomal copy number variations [31]. This in turn increased the resolution of our analysis, resulting in 24489 malignant epithelial cells extracted from tumor samples (Supplementary figure 1).

**Figure 1.**
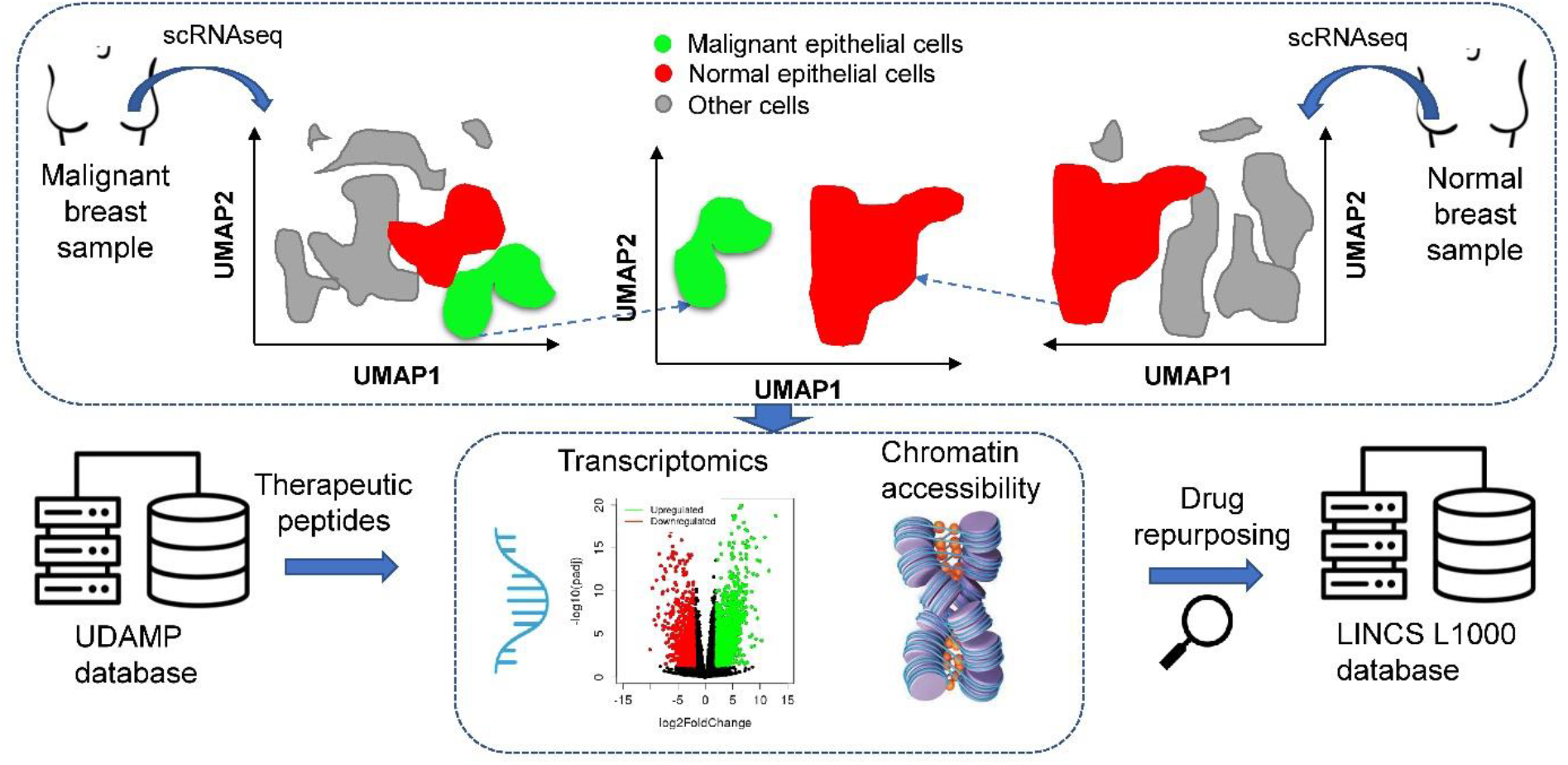
scRNA-seq provides single-cell resolution to compare different cell states. Using scRNA-seq, it is possible to extract the desired cell types in condition A (top left panel: malignant epithelial cells) and compare them to the same or different cell types in condition B (top right panel: nonmalignant epithelial cells). We studied the deregulation of endogenous therapeutic peptides derived from the University of Debrecen Antimicrobial and Immunomodulatory Peptide (UDAMP) database at different omics levels in BC epithelial cells. Accordingly, we investigated the available perturbagens in the Library of Integrated Network-based Cellular Signatures (LINCS) database and proposed candidate drugs that can change the expression of these peptides to eliminate the malignancy.

To acquire the normal epithelial cells, first we classified all cells using an unsupervised clustering method (see material and methods) (Figure 2A). Next, using Human Primary Cell Atlas (HPCA) [25] and Blueprint Epigenomics (BP) [26] as two previously annotated cell references, we could investigate the distribution of label scores among cells, (Figure 2B). We observed distinct scoring profiles among cells, demonstrating unambiguous concordance between cells and reference annotations and allowing us to accordingly assign labels to cells. In order to investigate the probable outliers in each labeled cluster, we examined the delta values for each cell, i.e., the difference between the score for the assigned label and the median across all labels for each cell. Few outliers were observed and the label assignments were deemed to be accurate (Supplementary Figure 2). Afterwards, to compare the concordance between the two independent methods of unsupervised clustering (figure 2A) and cell label assignment (figure 2B), we investigated the distribution of cell labels in each cluster. We observed strong agreement between the two methods (Figure 2C). Accordingly, we assigned the determined annotations to clusters (Figure 2D) and subsequently extracted the 21698 normal epithelial cells for downstream analysis.

**Figure 2.**
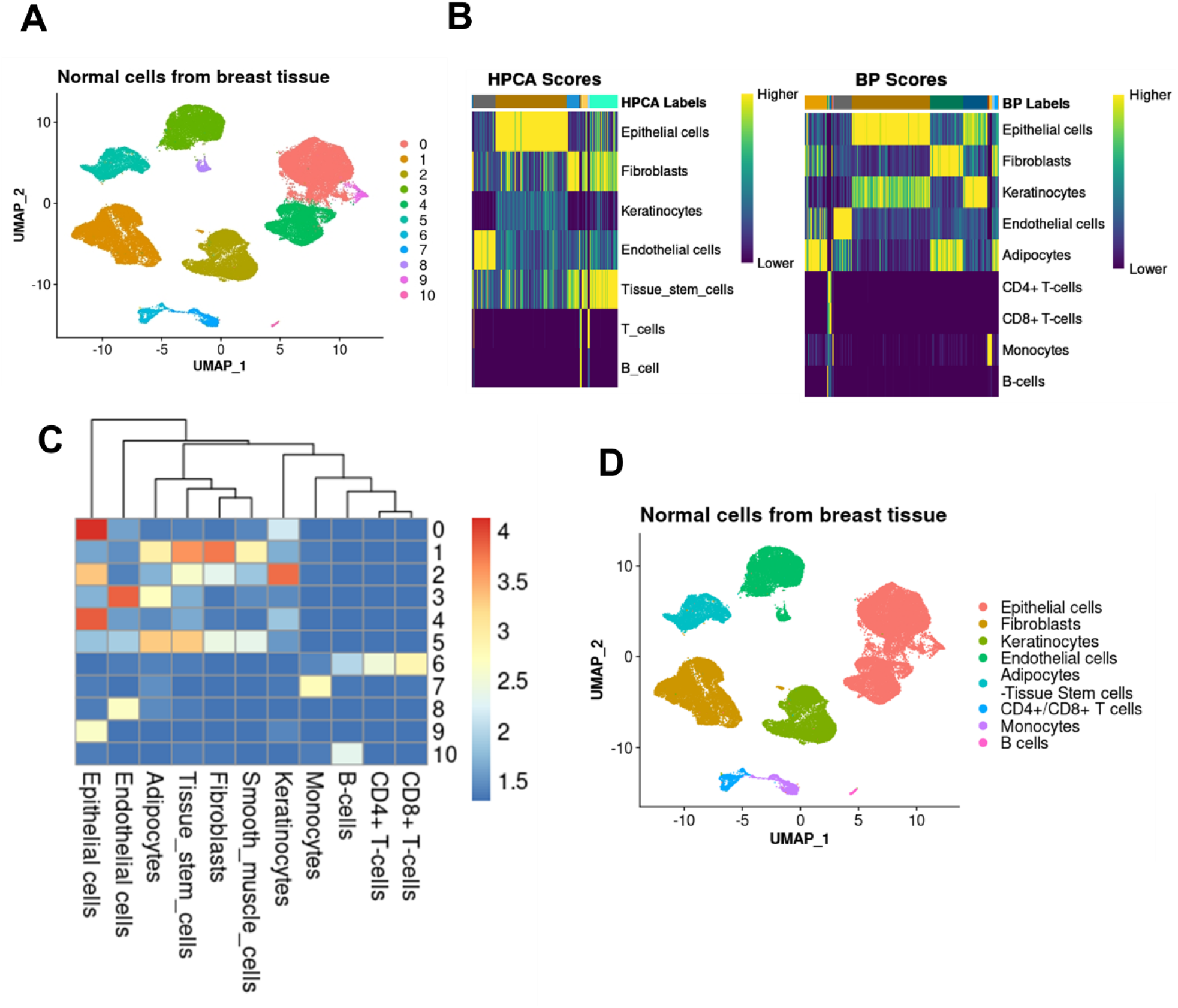
Using a combination of previously labeled datasets as a reference, automatic annotation can define cell types in scRNA-seq analysis. **A)** Unsupervised clustering of the cells based on the expression values of highly variable genes. **B)** Heatmaps of a nested matrix of per-cell scores showing correlation-based scores, prior to any fine-tuning, for each cell (rows), reference label color (columns) and reference label assignment as row names. **C)** Investigating the distribution of assigned cell labels in Panel B to the clusters defined in Panel A. Higher values indicate stronger concordance between two methods. Row numbers represent the cluster number in panel A. **D**) Assignment of labels deduced in Panel B to clusters defined in Panel A.

### Differential expression analysis to track APMs

For joint analysis of data from multiple scRNA-seq datasets (e.g., differential gene expression analysis between malignant and normal epithelial cells), we need to harmonize them into a single reference prior to any comparisons in order to remove technical artifacts. To do so, we used scRNA-seq data integration [27]. We identified cell pairwise correspondences between single cells in two sets, henceforth termed “anchors”. Anchors represent the cellular relationships across datasets, which can be used for scRNA-seq data integration by recovering the matching cell states (See material and methods) [27].

After data integration (Figure 3A), we performed differential gene expression (DEG) analysis (BC subtypes vs normal) to explore upregulated genes encoding immunomodulatory peptides in BC (Figure 3B). AMPs were retrieved from UDAMP [6] (Supplementary Data 1). Ten immunomodulatory peptides were selected for downstream analysis. These ten AMPs were either upregulated or differentially expressed in all BC subtypes. Among the selected AMPs, only in the ER BC subtype, S100A8 and S100A9 genes were downregulated.

**Figure 3.**
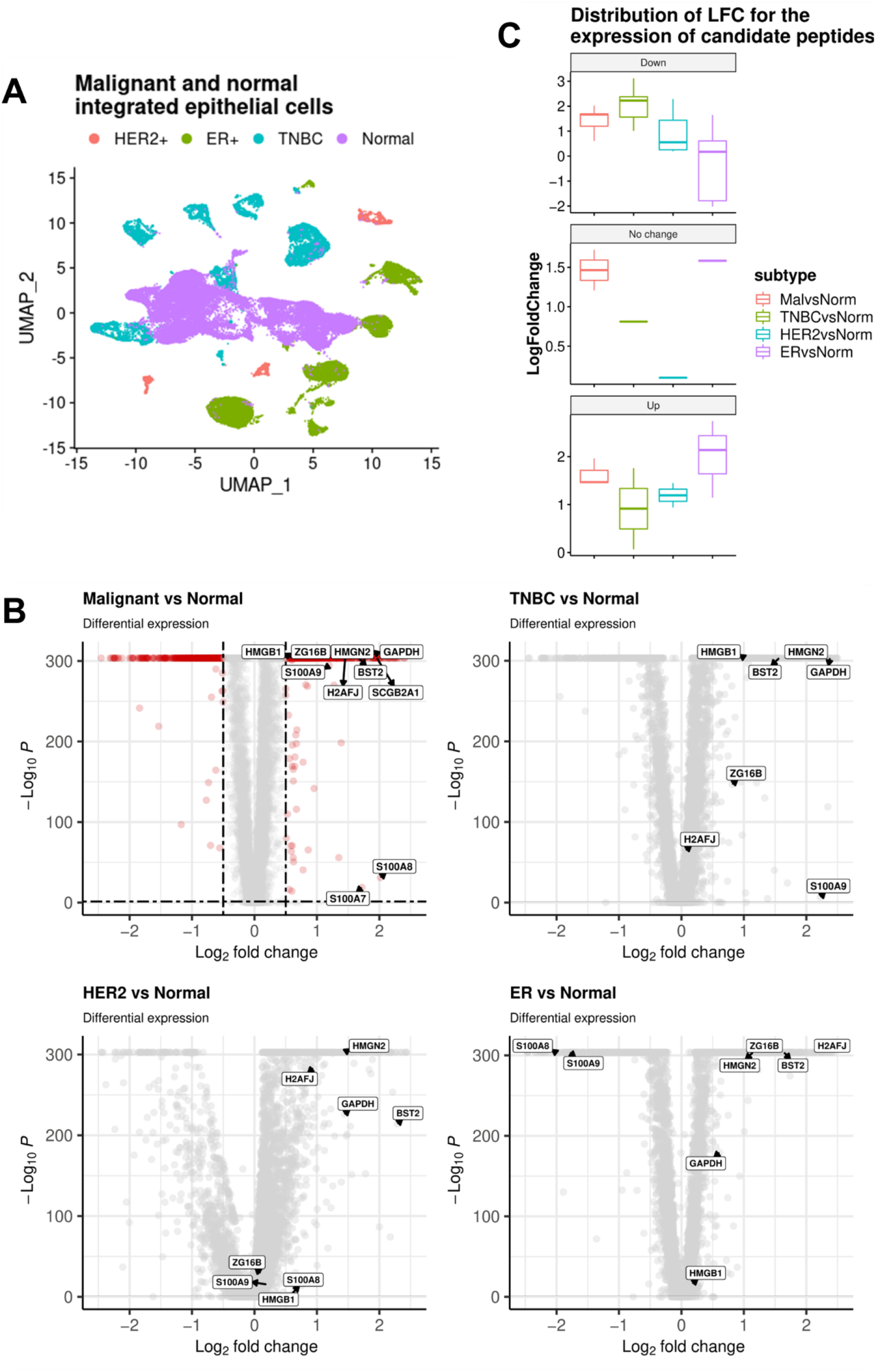
scRNA-seq data integration can correct the technical differences between datasets for accurate joint analysis. **A)** Malignant epithelial cells from three breast cancer subtypes were integrated with normal epithelial cells to remove the non-biological differences. **B)** Differential gene expression analysis shows ten immunomodulatory peptides being upregulated in malignant versus normal epithelial cells. Out of these ten, two are downregulated in ER BC subtype. **C)** According to the role of upregulated immunomodulatory peptides in cancer biology, three drug repositioning strategies were assigned to them; Downregulation, if they should be downregulated; Upregulation, if the expression should be enhanced; No change, if they are marker genes or have dual behavior in cancer biology (Table 1). Taking this classification into account, the distribution of LFC for different comparisons between BC subtypes and normal epithelial cells is illustrated.

The selected AMPs are a part of the innate immune system with a therapeutic effect against pathogens (Table 1). However, their role as immunomodulatory peptides against cancer is variable. We categorized these ten AMPs into three groups based on their association in cancer biology. The first group contains the genes that are connected with invasiveness, poor survival, disease severity, cancer cell proliferation and metastasis. These genes were grouped as “Downregulation”, as we aimed to decrease the expression of these genes using drug repositioning. The second group contains the genes associated with less aggressive behavior, more favorable outcomes in cancer and relapse-free survival. These genes were classified as “Upregulation” since we attempted to elevate their expression using drug repositioning. The last group was named “No change” as it includes cancer markers or the genes that show dual behavior in different malignancies. This group contains the genes that we did not consider for drug repositioning. We assessed the distribution of the expression of the genes encoding these peptides by repositioning strategy (figure 3C). Accordingly, TNBC had the highest and lowest LFC for the genes that should be down- and upregulated respectively through drug repositioning. It might be connected to the fact that TNBC is the most invasive BC subtype [32]. Hence, it is overexpressing the AMPs that are connected with worth prognosis and should be downregulated and less expression of the ones that are associate with favorable outcomes and should be upregulated.

**Table 1.**
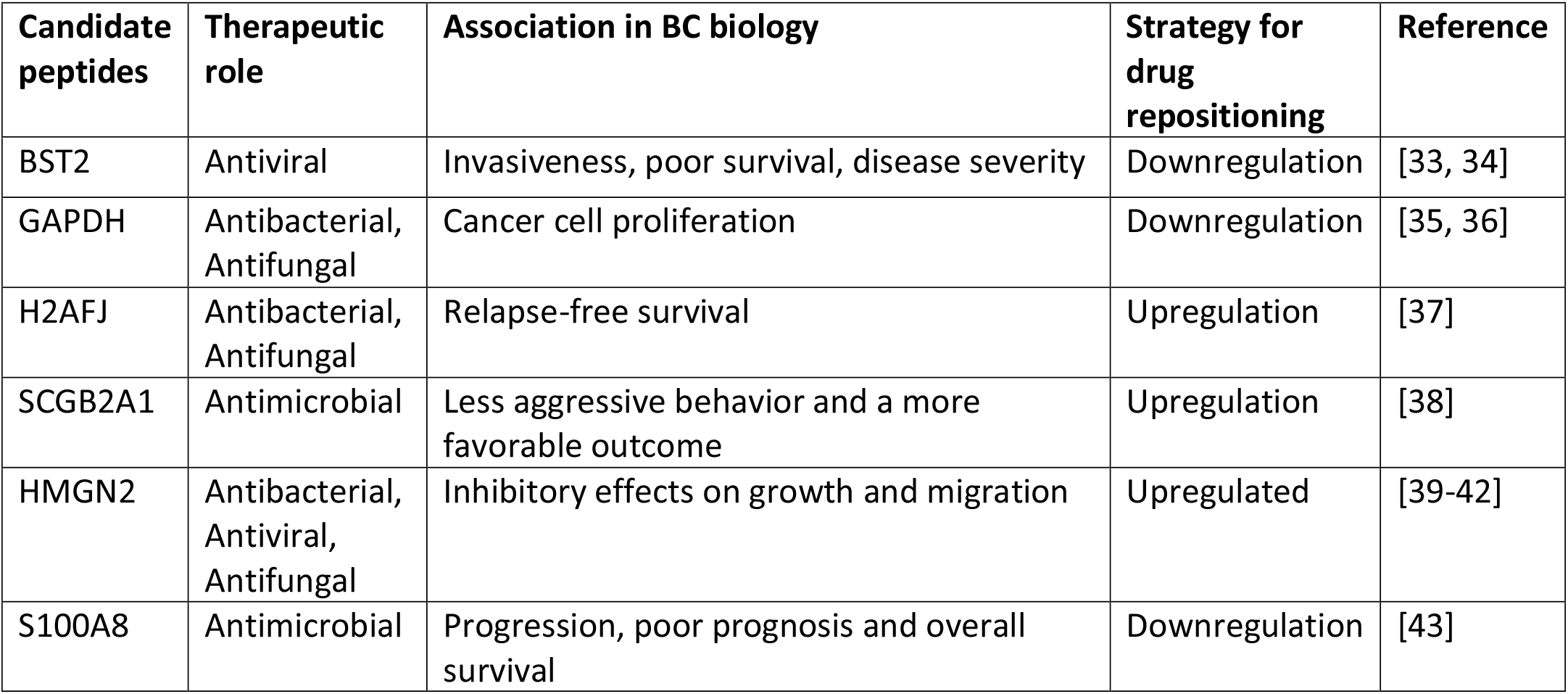
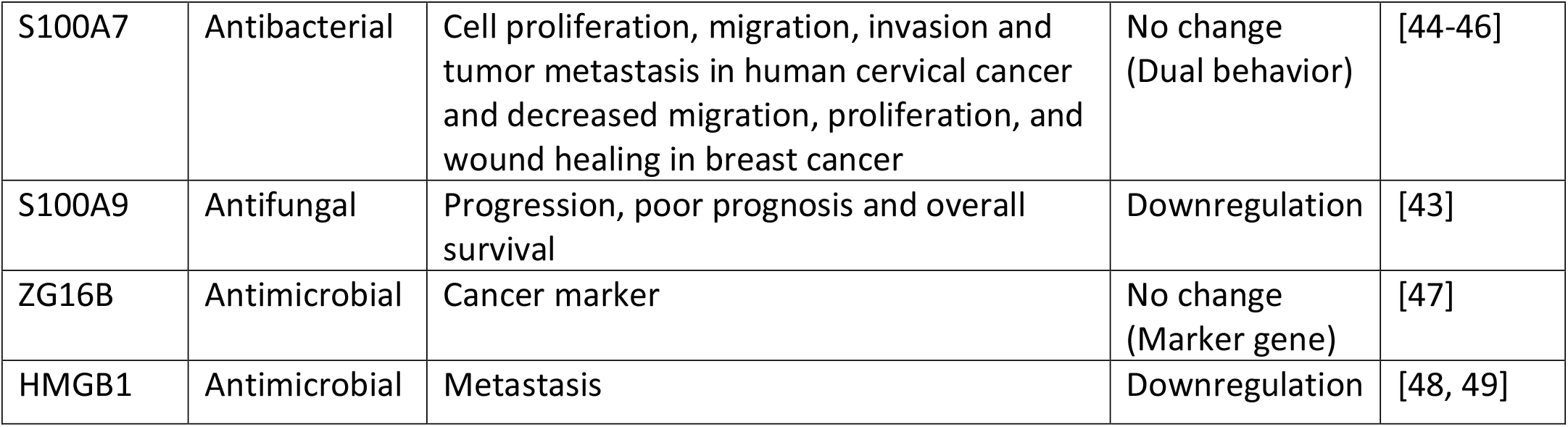
Deregulation of immunomodulatory peptides can be an immunotherapy strategy. Based on the association of ten upregulated immunomodulatory peptides in cancer biology, we determined a strategy for drug repositioning.

| Candidate peptides | Therapeutic role | Association in BC biology | Strategy for drug repositioning | Reference |
| --- | --- | --- | --- | --- |
| BST2 | Antiviral | Invasiveness, poor survival, disease severity | Downregulation | [33, 34] |
| GAPDH | Antibacterial, Antifungal | Cancer cell proliferation | Downregulation | [35, 36] |
| H2AFJ | Antibacterial, Antifungal | Relapse-free survival | Upregulation | [37] |
| SCGB2A1 | Antimicrobial | Less aggressive behavior and a more favorable outcome | Upregulation | [38] |
| HMGN2 | Antibacterial, Antiviral, Antifungal | Inhibitory effects on growth and migration | Upregulated | [39-42] |
| S100A8 | Antimicrobial | Progression, poor prognosis and overall survival | Downregulation | [43] |
| S100A7 | Antibacterial | Cell proliferation, migration, invasion and tumor metastasis in human cervical cancer and decreased migration, proliferation, and wound healing in breast cancer | No change (Dual behavior) | [44-46] |
| S100A9 | Antifungal | Progression, poor prognosis and overall survival | Downregulation | [43] |
| ZG16B | Antimicrobial | Cancer marker | No change (Marker gene) | [47] |
| HMGB1 | Antimicrobial | Metastasis | Downregulation | [48, 49] |

In order to increase the accuracy of our analysis, we repeated annotation, data integration, and DEG after an alternative normalization method, SCTransform [28]. We showed the compatibility of the results in both approaches. The same set of AMPs was upregulated in both methods and the same pattern of deregulation for candidate AMPs in ER BC subtype was observed (Supplementary figure 3).

### Spatial distribution of candidate genes

In order to investigate the spatial distribution of candidate genes, we extracted four ST datasets from the study conducted by Wu et al. (2021) [1]. To define the cell type composition of ST spots in these four ST datasets, we used scRNA-seq dataset derived from the same patients [1]. We performed the spot deconvolution using scRNA-seq and ST data integration. All the spots which were occupied by epithelial cells were labeled as malignant except the tissue section from CID44971 patient with some spots labeled as normal epithelial cells (Figure 4A). We calculated the expression scores of candidate genes across ST cell type clusters in all four sets. Except “HMGN2”, the rest of the candidate genes were significantly overexpressed in malignant compared to normal epithelial cells (Figure 4B). We performed differential gene expression analysis across all four tissue sections between spots which were labeled as malignant and normal epithelial cells. As a support for Figure 4B, “HMGN2” was the only downregulated gene in malignant epithelial cells while all the other gene candidates were significantly (p<0.05) upregulated (Figure 4C).

**Figure 4.**
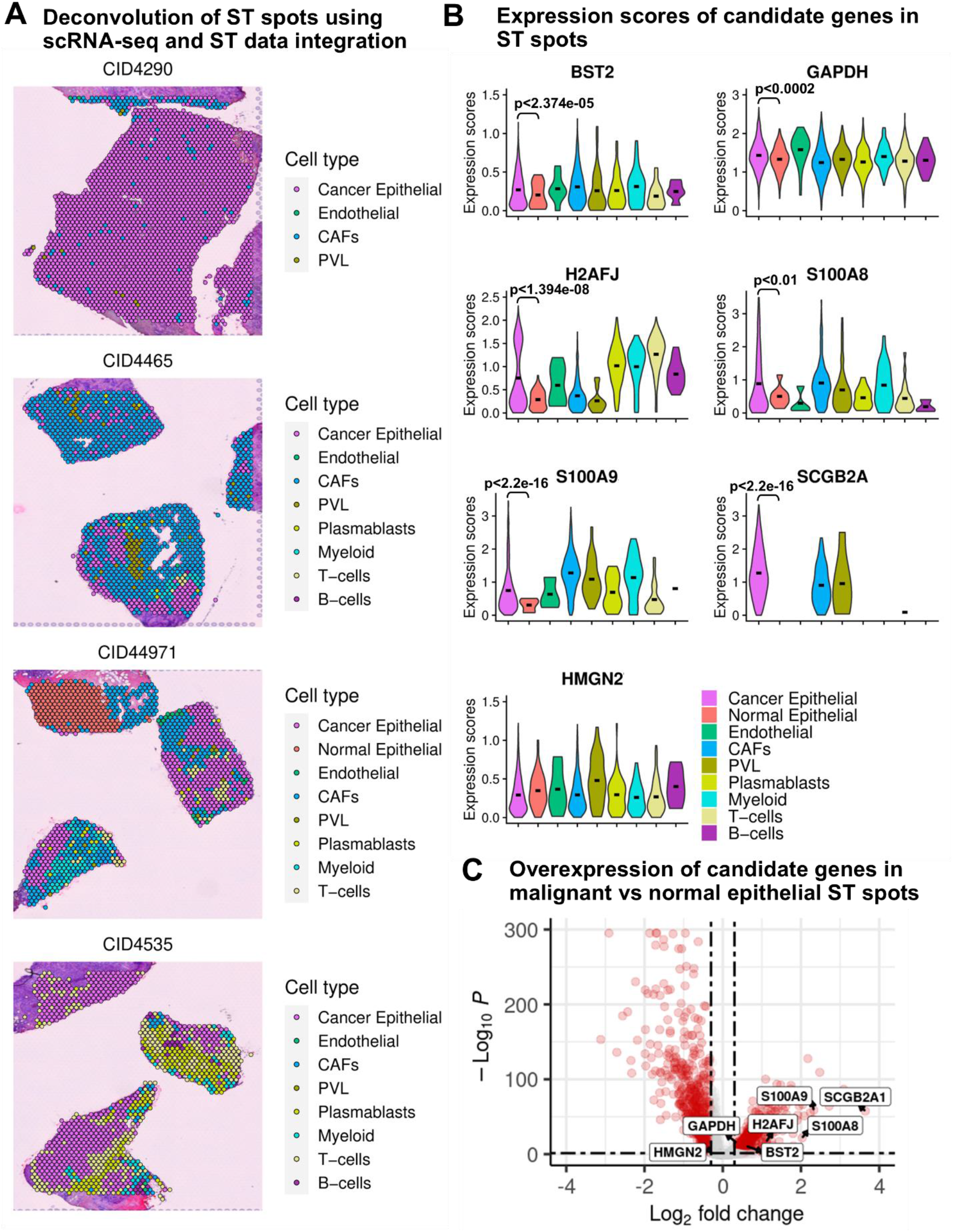
Investigation of the spatial distribution of candidate genes shows higher expression in the regions that are assigned to malignant epithelial cells when compared to locations with normal epitheial cells. **A)** Deconvolution of ST spots using data integration of four pairs of scRNA-seq and ST datasets from the same patients. **B)** Comparing the expression scores of candidate genes across spots in all four tissue sections which their cell type composition is unveiled using deconvolution methods. **C)** Differential gene expression analysis between ST spots assigned to malignant epithelial cells and normal epithelial cells in all four tissue sections.

### Exploring the status of candidate APMs in chromatin and gene levels

To validate our findings from scRNA-seq data at the chromatin level, we obtained ATAC-seq data for four different BC epithelial cell lines (two TNBC cell lines with repetition [23], one ER BC cell line with repetition [24] and one normal [50] breast epithelial cell line). The peaks show higher accessibility of chromatin for candidate AMPs cancerous cells versus normal (Figure 5A). This is consistent with the results from scRNA-seq which shows the upregulation of these AMPs in BC versus normal epithelial cells (Figure 3B). Hence, the ATAC-seq data supports results from scRNA-seq analysis.

**Figure 5.**
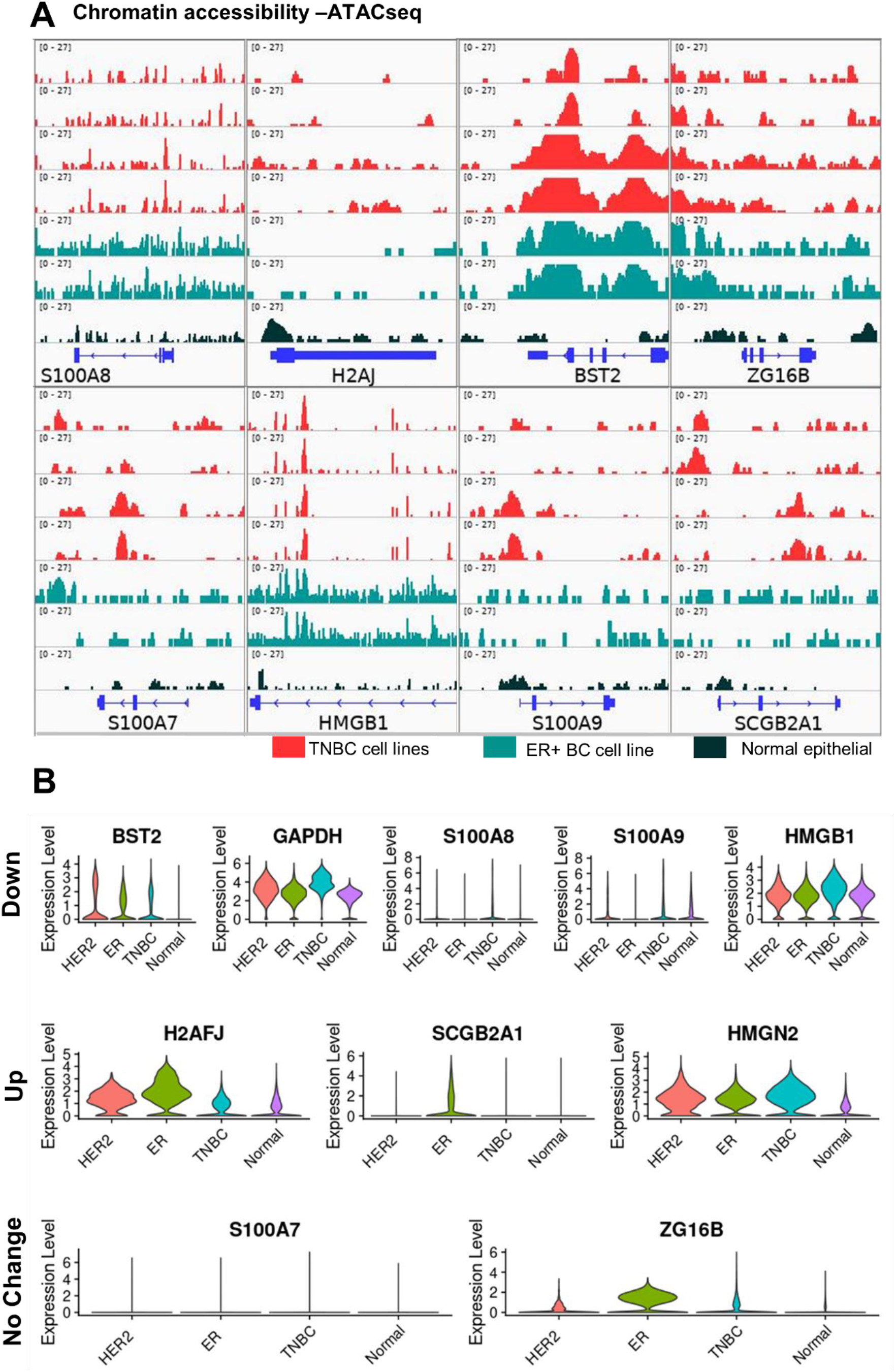
Collecting the information from different omics levels increases the accuracy of data analysis. **A)** Chromatin accessibility data from two TNBC cell lines with a repetition (MDA-MB-231, MDA-MB-436), one ER cancer cell line with a repetition (MCF7) and a normal epithelial cell (MDF10A) is shown. Chromatin is more accessible in malignant cells compared to normal for candidate genes. **B)** Distribution of expression of candidate genes are separated based on drug repositioning strategies.TNBC has the highest expression of the genes that should be downregulated.

In gene level, distribution of expression of candidate genes by drug repositioning strategy (Figure 5B) was consistent with differential expression (Figure 3B). All three BC subtypes had higher expression of selected AMPs in comparison with normal epithelial cells. TNBC had the highest expression of the genes belonging to downregulation drug repositioning strategy. Similarly, ER BC had the lowest and mostly highest expression of genes belonging to up- and downregulation strategies, respectively. Hence, TNBC appears to be the first concern among all BC subtypes studied which might be connected with the higher invasiveness of TNBC versus ER and HER2 BC subtypes [32].

### Drug repositioning

Our goal of drug repositioning is to deregulate the candidate AMPs based on their role in cancer biology. Hence, we need to know which pathways we may intervene during this process. In order to study the functional aspects of candidate APMs, we performed a correlation followed by Gene Ontology (GO) and KEGG pathway analysis. The correlation was calculated among the top 6000 variable genes in the integrated scRNA-seq dataset (Figure 3A). The top 20 significantly correlated genes (if available) with candidate AMPs were extracted and applied to GO and KEGG pathway analysis. We found that BST2 participates in the MHC protein complex according to GO (Supplementary figure 4A), and based on KEGG, it is connected with antigen processing and presentation (Supplementary figure 4B). These enriched terms are due to the correlation of BST2 with B2M, HLA-B, and HLA-C genes (Supplementary figure 4C). Thus, we need to be cautious about downregulating BST2 in a way that repurposed drugs only target BST2 and not the correlated MHC genes. In addition, S100A7, S100A8 and S100A9 AMPs are correlated with genes in the IL-17 signaling pathway (Supplementary figure 4B). It has been shown that IL-17 enhances inflammation and metastasis in BC [51]. S100A8 and S100A9 were classified as genes belonging to the downregulation strategy by drug repositioning while S100A7 was not a target for drug repurposing due to its dual behavior in different cancers [44-46]. Besides, HMGN2, a gene in the drug repositioning upregulation strategy group, is correlated with the p53 signaling pathway (Supplementary figure 4B). In general, exposure of cells to different stress signals (e.g. malignancy) activates the p53 signaling pathway. Consequently, cells activate several transcriptional programs including cell cycle arrest, DNA repair, senescence, and apoptosis leading to the suppression of tumor growth [52].

Taking the amino acid content of candidate AMPs into account, we examined their potency to display anticancer properties. According to the high specificity, strong tumor penetration capacity, and low toxicity to normal cells, anticancer peptides are considered as an alternative to chemotherapy drugs [53]. Hence, we used the iDACP database (http://mer.hc.mmh.org.tw/iDACP/) [53] to predict the anticancer potential of the candidate genes. We found that H2AFJ was predicted to be a potential anticancer peptide with the maximum available prediction specificity in the database (90%). Interestingly, H2AFJ is among the AMPs that were planned to be upregulated through drug repositioning. So, H2AFJ not only is associated with relapse-free survival [37], but directly may eliminate the cancer cells.

To explore the effect of previously approved drugs against candidate AMPs in BC, we extracted all available perturbagen data for all BC cell lines in the LINCS L1000 database. LINCS L1000 is an actively growing collection of gene expression profiles for thousands of perturbagens at a variety of time points, doses, and cell lines [54]. Among the BC cell lines in LINCS L1000, we chose MCF7 for downstream analysis. The rationale behind our selection was the number of available perturbagens for each BC cell line. According to the data availability in LINCS L1000, among the BC cell lines, the number of perturbagens for MCF7 is considerably higher than other cells (Supplementary figure 5A). Hence, it provides a great opportunity to discover potential drugs out of ∼30000 perturbagens that can be repurposed in our study.

We computed the LFC values for all MCF7 drug perturbagens versus DMSO as control (see material and methods). Next we extracted the LFC values of the candidate AMPs across all ∼30000 perturbagens (Supplementary Data 2 and 3). According to the drug repositioning strategy and immunomodulatory peptide gene deregulation by drugs, we selected the top 20 perturbagens that can up or downregulate the candidate AMPs. We investigated the effect of selected drugs (Supplementary Data 3) across all selected AMPs. To make sure that downregulation of BST2 does not decrease the expression of its correlated genes which encode the MHC protein complex, we included those genes (i.e, B2M, HLA-B, and HLA-C genes) in our analysis. Hence, we showed that the candidate drugs does not intervene in the expression of the other AMPs and only affect their own target (Figure 6A, Supplementary figure 5B). Thus, they can efficiently and specifically deregulate the AMPs based on the drug repositioning strategy. Moreover, the candidate drugs against BST2 does not affect the expression of its correlated genes which enroll in the MHC complex. All deregulations in Figure 6A are compatible with drug repositioning strategy in Table 1.

**Figure 6.**
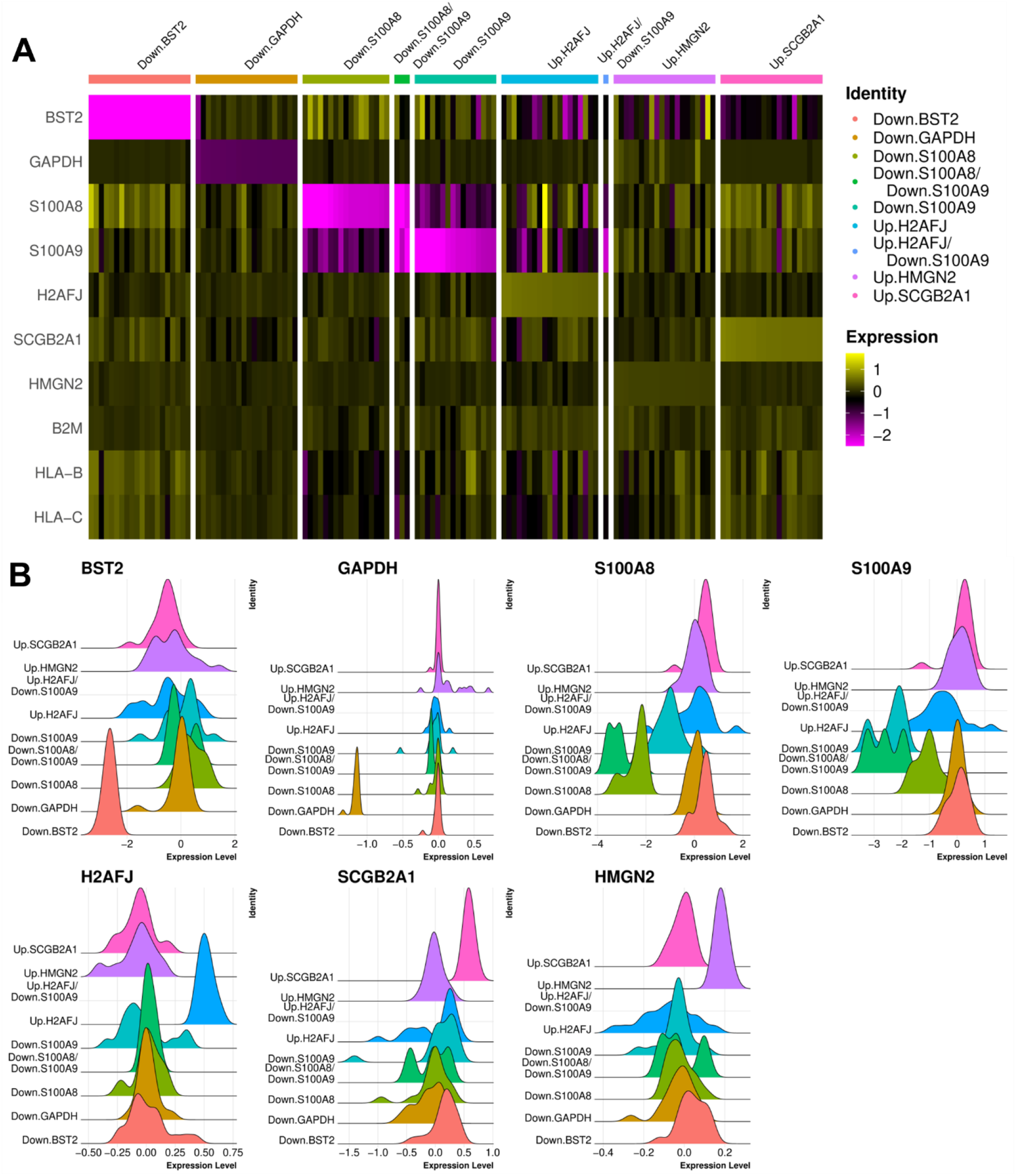
Drug repositioning can find new indications for previously approved drugs. **A)** Drugs as column names of the heatmap are separated into nine groups. The name of each group starts with the drug repositioning strategy and the target gene (i.e Down: repurposed drugs aim to downregulate the gene; Up: repurposed drugs aim to upregulate the gene). Results from the heatmap show high concordance between defined strategy and the effect of drugs on candidate genes. **B)** Categorization of the drugs in the Y axis is similar to Panel A. Deregulation of the distribution of LFCs for candidate genes matches the strategy for repurposed drugs.

In addition, we investigated the distribution of LFC values across all cells to validate the specificity of the candidate drugs. Accordingly, based on the intensity of changes in gene expression by drugs (Figure 6B and Supplementary Figure 5B) and changes in average and percentage of LFC values across all candidate genes (Supplementary Figure 5C) we showed that repurposed drugs only decrease or increase the expression level of target gene. Some of the drugs were common between selected perturbagens against different AMPs. Thus, they are multi-functional in our study. For instance, some of the proposed drugs can simultaneously decrease the expression of S100A8 and S100A9 (Figure 6A and B, Supplementary Figure 5B and C). Besides, a group of drugs increase the expression of H2AFJ and decrease the expression of S100A9, at the same time. Their effect is not visible in figure 6B and Supplementary figure 6B due to the low number of them in the dataset (Supplementary Data 2). However, their effect is illustrated in Supplementary figure 6C. Thus, we showed the concordance of changes and the drug repositioning strategy, drug repurposing specificity and efficiency.

## Discussion

BC arises from breast epithelial cells that acquire specific genetic variations leading to the subsequent loss of tissue homeostasis [55]. However, not all of the epithelial cells are malignant in a cancerous tissue [1]. To raise the accuracy of our analysis, we extracted only the transcriptomics profiles of malignant epithelial cells in cancer patients and subsequently compared them to normal epithelial cells. The same approach has been frequently used in different cancer studies using InferCNV method [20, 56, 57]. This gave us the opportunity to investigate the changes in the expression of, to the best of our knowledge, all so far known human endogenous AMPs [6] in malignant and nonmalignant breast epithelial cells.

We showed the overexpression of ten immunomodulatory peptides in gene level of BC epithelial cells versus normal and validated these results in chromatin level. The elevated expression of these AMPs in BC is also reported previously [34, 38, 39, 43, 44, 47, 48, 58, 59]. However, the therapetuic use of these peptide against BC by manipulationg their expression is yet to be explored.

Pavlicevic et al. [19] introduced the AMPs as a promising source for drug discovery. These components are a complex class of bioactive peptides with diverse effects on both innate and adaptive immunities. The main focus of Kumar et al. [6] on data curation of AMPs in UDAMP database was on their antimicrobial, antifungal and antiviral features. Hence, we used this source to study the elevated immunomodulatory peptides in BC to see if they are potential to be anticancer, cancer driver or ineffective in cancer progression (Table 1). AMPs can have one or a combination of roles [6, 60]. We observed that a conflict in these roles may appear. For instance, an antimicrobial peptide may promote the cell growth of malignant cells (Table 1). Taking BST2, GAPDH, S100A8, S100A9 and HMGB1 as examples, in spite of their advantageous role in different infections (Supplementary Data 1), they contribute to the cancer cell proliferation, invasiveness, poor survival, and disease severity in different cancers [33-36, 43, 48, 49]. In addition, S100A7 has a dual behavior in different malignancies. In cervical cancer and BC [44-46] S100A7 leads to increased and decreased cancer cell proliferation, respectively, while it is an antibacterial peptide [6]. Hence, this is necessary to understand how the upregulated immunomodulatory peptides in breast malignant epithelial cells act and how we can deregulate these peptides in the favor of patients. One of the ideal approaches for deregulation of target genes is using previously approved drugs for the new indication termed “drug repositioning.”

The use of LINCS L1000 project [54] in drug repositioning for immunotherapy has been already addressed [61, 62]. Besides, several studies attempt to repurpose the non-human immunomodulatory peptides [63] and immunomodulatory drugs against different disorders [64]. But, this is the first research which focuses on repurposing the drugs in order to modulate the expression of human immunomodulatory peptides to eliminate the BC. By categorizing the upregulated AMPs in BC vs normal epithelial cells into the anticancer, cancer driver and ineffective in cancer progression, we investigated the drugs that can deregulate them in favor of patients. Before proceeding to drug repositioning, we checked the gene-gene correlation and functional analysis of upregulated AMPs in BC to figure out which pathways we may intervene. Among all candidate AMPs (Table 1), the correlated genes with BST2 are connected with MHC complex [65]. So, we tracked only the drugs that deregulate BST2 and not its correlated genes including B2M, HLA-B and HLA-C (Figure 6A, Supplementary Figure 4C). As a result, we have provided a list of drugs (Supplementary Data 3) that could potentially downregulate and upregulate the AMPs that act as cancer driver or anticancer in BC patients, respectively.

The anticancer effect of many drugs in Supplementary Data 3 have been already verified against BC. For instance, Mitoxantrone is connected with tumor-specific CD8+ T-cell response and anti-tumor activity [66] and the auxiliary treatment of Ivermectin and anti-PD1 antibody merits clinical testing in breast cancer [67]. In addition, Mepivacaine inhibited breast cancer cell viability and induced cytotoxicity [68] and Verteporfin inhibits cell proliferation and induces apoptosis in different subtypes of breast cancer cell lines [69]. Trifluoperazine hydrochloride suppresses triple-negative breast cancer tumor growth and brain metastasis [70] and Artemether increases the antitumor effect on breast cancer cells [71]. In conclusion, the safety and anticancer features of the proposed drugs have already been verified. However, their effect on deregulation of overexpressed AMPs in BC should be tested in further wet lab investigations. Having multi mode of actions can be a promising aspect for these drugs to eliminate the BC more efficiently.

## Data Availability

All data produced in the present work are contained in the manuscript

## Declaration

### Ethics approval and consent to participate

‘Not applicable’ for that section

### Consent for publication

‘Not applicable’ for that section

### Code availability

We uploaded the scripts in R programming language to GitHub: https://github.com/ElyasMo/Drug.rep_BC

### Availability of data and materials

The scRNA-seq data for breast cancer and normal patients are publicly available through the Gene Expression Omnibus under accession number GSE176078 and GSE161529, respectively. The ATAC-seq data for BC cell lines used in this study are publicly available through the Gene Expression Omnibus under accession number GSE114964 and GSE174152. The spatially resolved transcriptomics datasets used in this study are available from the Zenodo data repository (https://doi.org/10.5281/zenodo.4739739).

### Author contributions

Conceptualization: E.M., M.H.S., N.S., and S.D.; Methodology: E.M.; Software: E.M.; Validation: E.M.; Formal analysis: E.M.; Investigation: E.M., M.H.S., A.M., C.Z., N.S, and S.D.; Data Curation: E.M; Writing - Original Draft: E.M.; Writing - Review & Editing: E.M., C.Z., H.J., A.M., M.H.S., S.L, N.S. and S.D.; Visualization: E.M.; Supervision: M.H.S. and A.M., M.T.; Project administration: M.H.S.; Funding acquisition: M.H.S.

## Supplementary Figures

**Figure S1.**
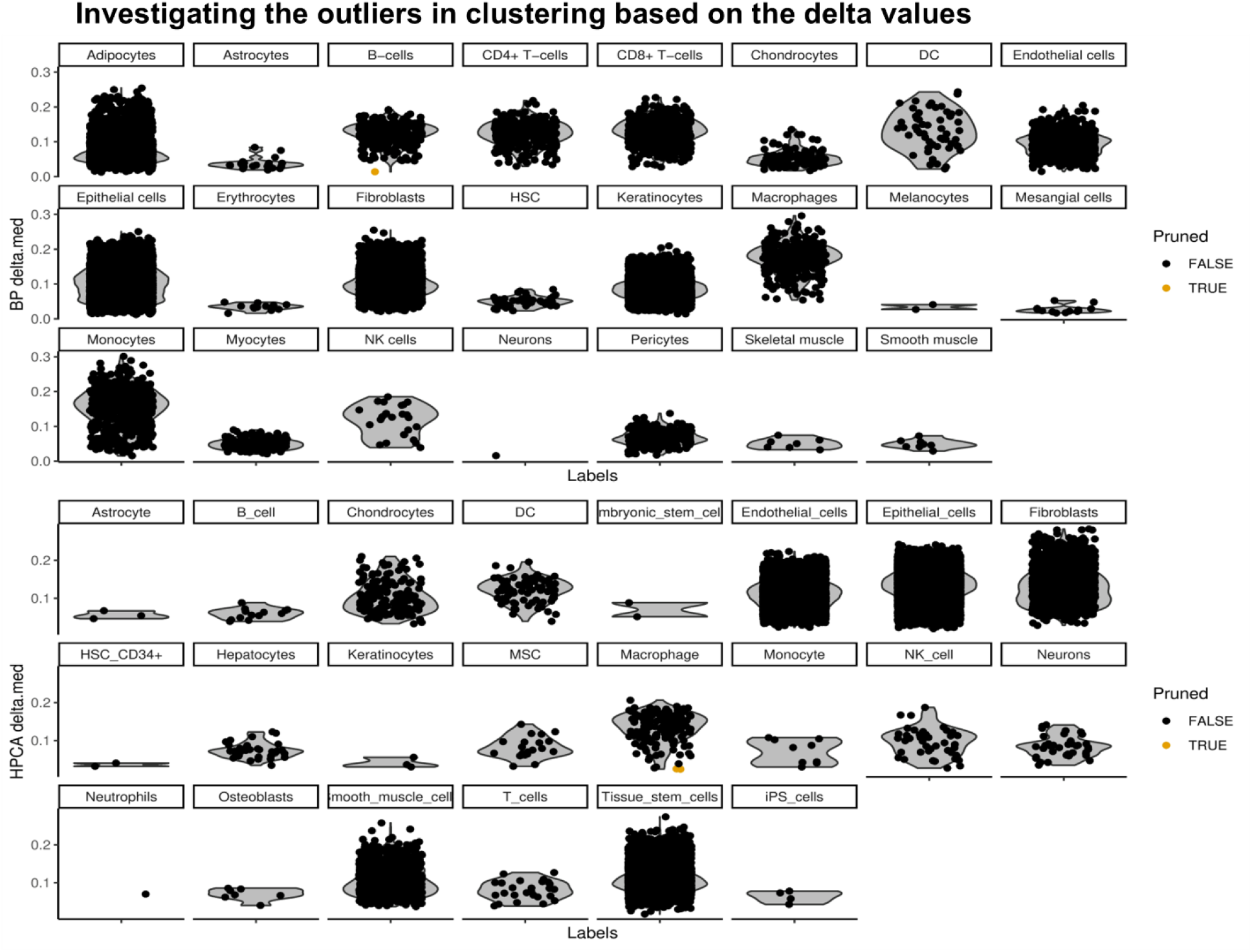
The accuracy of cell type annotation was approved by multiple tests. We measured the delta values for each cell in figure 2B, i.e., the difference between the score for the assigned label and the median across all labels for each cell. Outliers are shown in yellow.

**Figure S2.**
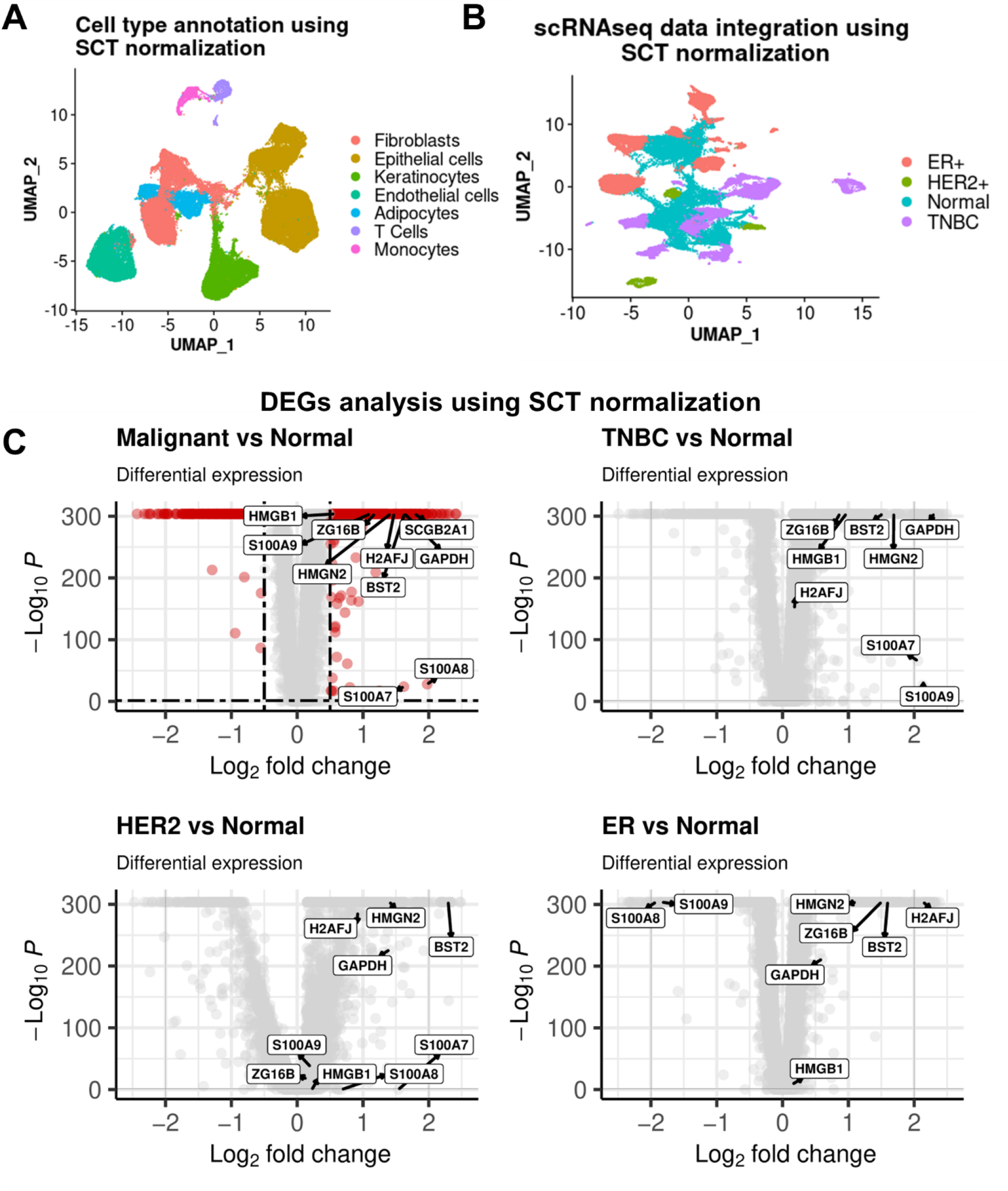
Normalization approach does not affect the obtained results in differential gene expression. **A)** We performed the cell type annotation using SCTransform data normalization. **B)** We did the data integration using the SCTransform approach. **C)** Results from differential gene expression analysis using the SCTransform method are compatible with our previous findings.

**Figure S3.**
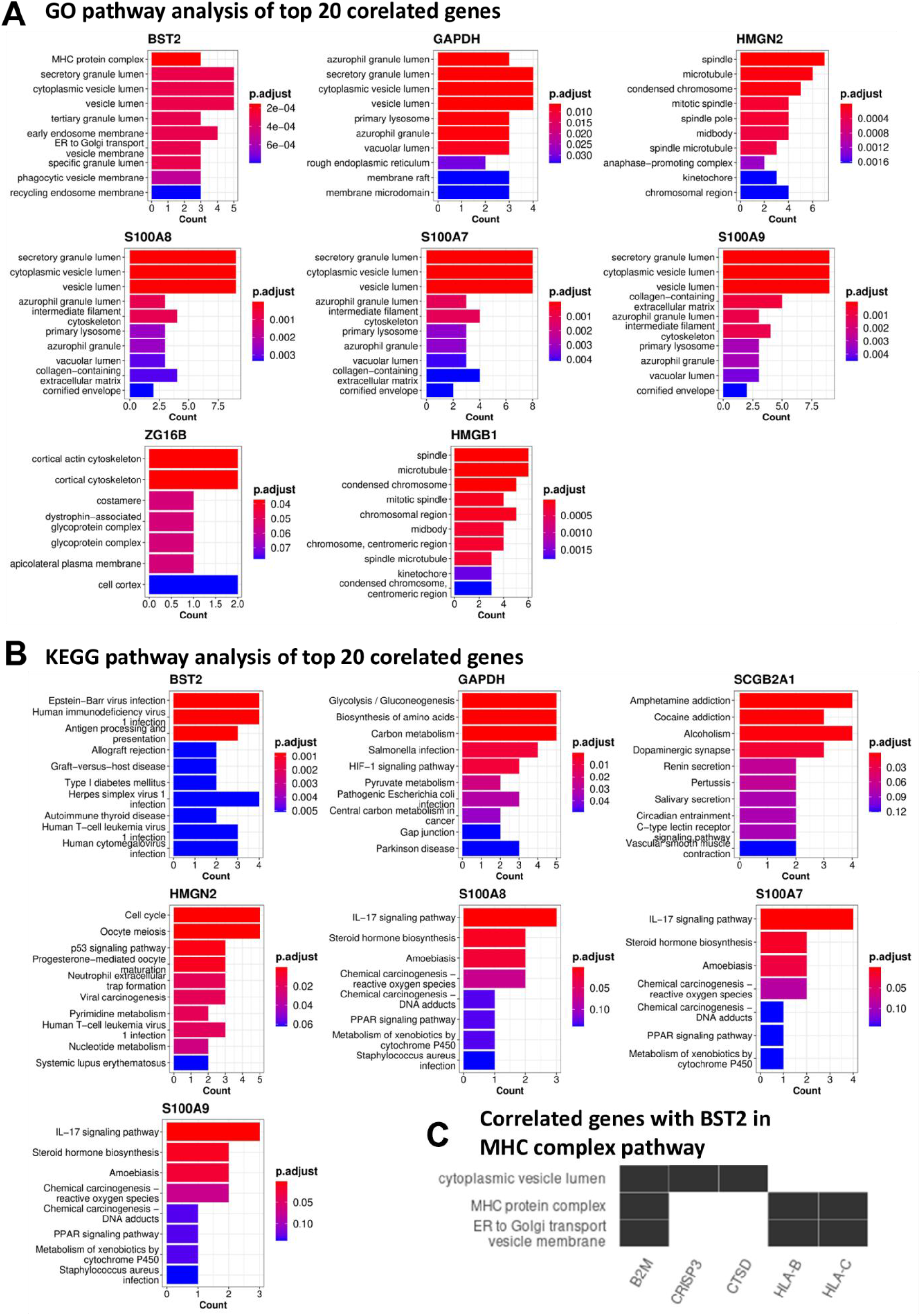
Functional analysis of candidate AMPs and their correlated genes can give an insight on their role in BC biology. **A)** Gene ontology pathway analysis of candidate immunomodulatory peptides and top 20 correlated genes. **B)** KEGG pathway analysis of candidate immunomodulatory peptides and top 20 correlated genes. **C)** Correlated genes with BST2 that take part in the MHC protein complex pathway.

**Fig S4.**
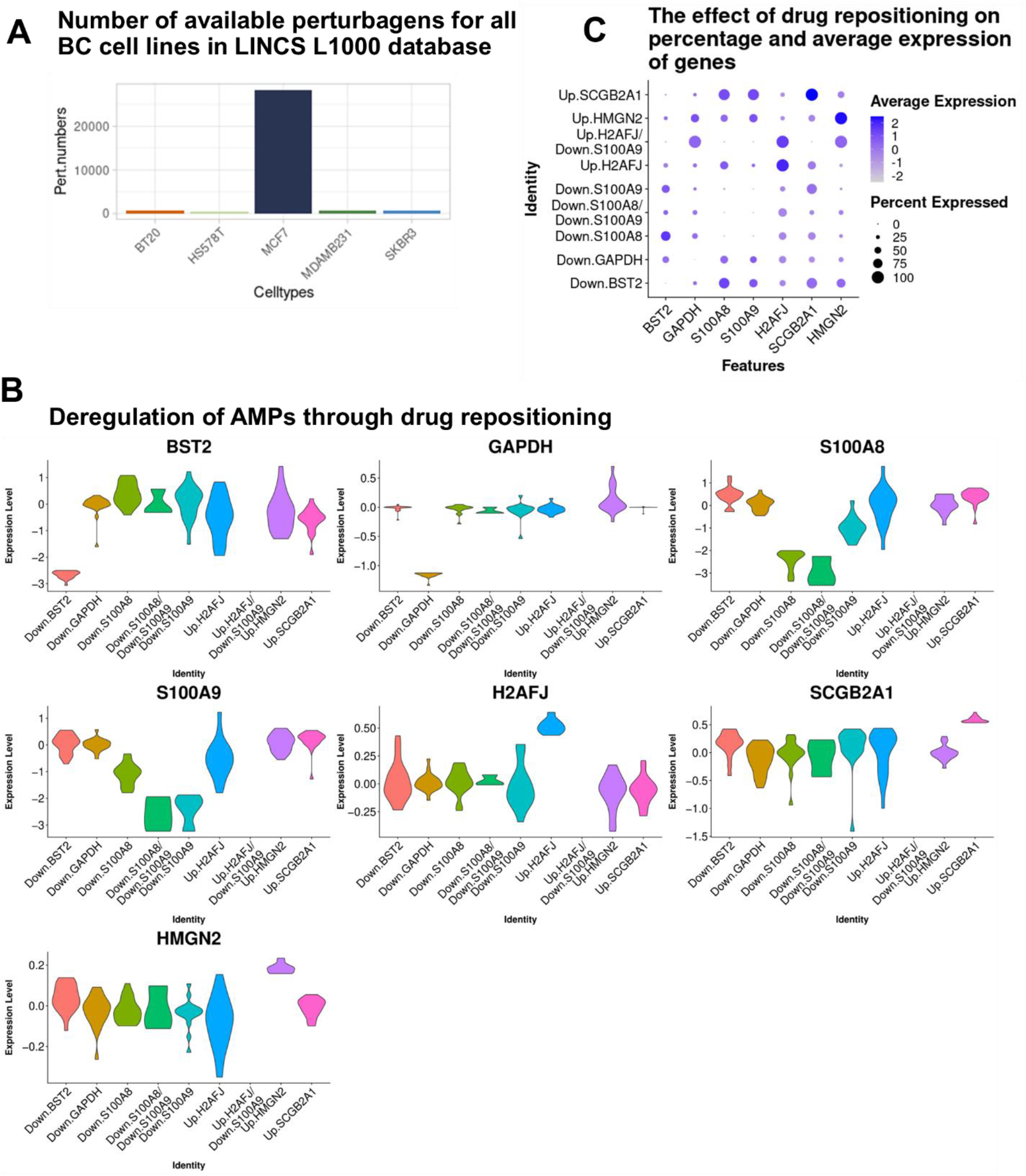
Distribution of expression of immunomodulatory peptides is purposefully deregulated by repurposed drugs. **A)** Number of available perturbagens for all BC cell lines in LINCS L1000 database. **B)** Deregulation of the distribution of LFCs for candidate genes matches the repurposed drugs strategy. Due to the specificity of the repurposed drugs, only the target gene(s) are deregulated based on the determined aim(s) of the drugs **C)** The changes in the percentage and average of LFC values of genes are compatible with the determined aim(s) of the drugs.

